# Diagnostic Accuracy of Carotid Plaque Instability by Noninvasive Imaging: A Systematic Review and Meta-analysis

**DOI:** 10.1101/2023.09.25.23296124

**Authors:** David Pakizer, Jiří Kozel, Patrick Taffé, Jolanda Elmers, Janusz Feber, Patrik Michel, David Školoudík, Gaia Sirimarco

**Author notes:** Corresponding author, Prof. David Školoudík, M.D., Ph.D., FESO, FEAN Centre for Health Research Faculty of Medicine, University of Ostrava Syllabova 19, CZ-703 00 Ostrava, Czech Republic. (= equal contribution) (David Školoudík, Gaia Sirimarco).

## Abstract

**Background:** There is increasing evidence that plaque instability in the extracranial carotid artery may lead to an increased stroke risk independently of the degree of stenosis. We aimed to determine diagnostic accuracy of vulnerable and stable plaque using noninvasive imaging modalities when compared to histology in patients with symptomatic and asymptomatic carotid atherosclerosis.

**Methods:** Medline Ovid, Embase, Cochrane Library, and Web of Science were searched for diagnostic accuracy of noninvasive imaging modalities (CT, MRI, US) in the detection of 1) vulnerable/stable plaque, and 2) vulnerable/stable plaque characteristics, compared to histology. The quality of included studies was assessed by QUADAS-2 and univariate and bivariate random-effect meta-analyses were performed.

**Results:** We included 36 vulnerable and 5 stable plaque studies in the meta-analysis, and out of 211 plaque characteristics from remaining studies, we classified 169 as vulnerable and 42 as stable characteristics (28 CT, 120 MRI, 104 US characteristics). We found that MRI had high accuracy (90% [95% CI:82–95%]) in the detection of vulnerable plaque, similar to CT (86% [95% CI:76–92%]; p>0.05), whereas US showed less accuracy (80% [95% CI:75– 84%]; p=0.013). CT showed a high diagnostic accuracy to visualize characteristics of vulnerable or stable plaques (89% and 90%) similar to MRI (86% and 89%; p>0.05); however US had lower accuracy (77%, p<0.001 and 82%, p>0.05).

**Conclusions:** CT and MRI have a similar, high performance to detect vulnerable carotid plaques, whereas US showed significantly less diagnostic accuracy. Moreover, MRI visualized all vulnerable plaque characteristics allowing for a better stroke risk assessment.

**Registration:** PROSPERO ID CRD42022329690 (https://www.crd.york.ac.uk/prospero/display_record.php?RecordID=329690)

## Introduction

Carotid artery atherosclerosis is one of the main causes of ischemic stroke in adults and also a marker of vascular health and for risk assessment.^1^ In addition to the degree of focal carotid stenosis, plaque components are important risk factors for plaque instability and subsequent risk of transient ischemic attack or stroke.

Vulnerable or unstable plaques are plaques with a high likelihood of rapid progression, and a substantial risk of ruptures subsequently leads to thrombus formation and symptoms.^2–5^ Therefore, detection of plaque characteristics may help to identify patients at high risk for cerebrovascular events.

In general, a vulnerable plaque is characterized by the presence of intraplaque hemorrhage (IPH), a thin or ruptured fibrous cap overlying a large lipid-rich necrotic core (LRNC), inflammation and neovascularization, and an ulcerated or fissured surface,.^6–8^ It has been demonstrated that an increasing size of LRNC and presence of thin/ruptured fibrous cap are strongly associated with risk of stroke.^9–10^ Moreover, the presence of IPH predicted plaque progression and cerebrovascular events in asymptomatic and symptomatic patients with a hazard ratio of 3.6 and of 4.8, respectively.^11–13^ Inflammation also plays an important part in the risk of plaque instability by causing structural changes in the vessel wall, destabilization and plaque rupture.^14^ On the other hand, fibrous tissue provides structural integrity of a stable carotid plaque and calcification is considered to be the main marker of stability, when lipids are either absent or present in only small amounts.^14–15^

Therefore, the performance of diagnostic imaging techniques to evaluate carotid plaques can have a major impact on risk assessment and management of patients. Noninvasive modalities such as ultrasound (US), computed tomography (CT) and magnetic resonance imaging (MRI) play a key role, because of their availability and high diagnostic accuracy.^16^

The aim of this study was to determine diagnostic accuracy of noninvasive imaging modalities (US, CT, MRI) for vulnerable and stable carotid atherosclerotic plaques compared to histology in adult patients with symptomatic and asymptomatic carotid plaques.

## Methods

PROSPERO (International Prospective Register of Systematic Reviews) was used for the prospective registration of our study (ID CRD42022329690).^17^ This systematic review and meta-analysis was performed according to the PRISMA (Preferred Reporting Items for Systematic Reviews and Meta-analyses) 2020 statement^18^ and the STARD (Standards for Reporting of Diagnostic Accuracy Studies) 2015 statement.^19^

### Study eligibility criteria

We included human studies that investigated atherosclerotic carotid artery disease *in vivo* using selected noninvasive modalities (US, CT or MRI as index test) to assess plaque composition compared to histology. Inclusion criteria were as follows: human subject research studies (retrospective or prospective); age of patients ≥ 18 years; studies evaluating patients with an atherosclerotic plaque in the cervical carotid arteries, including the extracranial internal and/or common carotid artery; studies using US, CT or MRI at the cervical carotid arteries to assess specific plaque features; studies that investigated correlations of the characteristics of the index plaque with histological analysis of the plaque (specimens were obtained from carotid endarterectomy or autopsy); reported number of true positive (TP), false positive (FP), false negative (FN), and true negative (TN) results for the diagnosis of any plaque characteristic, and available data to calculate sensitivity and specificity. Histology was used as the reference standard for the evaluation of plaque composition. Studies with a time window > 4 weeks (1 month) between carotid plaque imaging and histological assessment were excluded.

### Database searching

A systematic search strategy to find eligible studies was designed by a medical librarian (J.E.), using a combination of controlled vocabulary terms and free text terms covering the four overarching concepts of the study: “atherosclerotic carotid artery disease” AND “atherosclerotic plaque” AND “Imaging” AND “histology”. The search strategy was translated for the following databases: Medline Ovid ALL, Embase.com, Central – Cochrane Library Wiley, and Web of Science Core collection. A second medical librarian peer-reviewed all search equations before the searches were conducted (27/06/2022). Searches were performed without limits for publication date or language and the search strategies and search engines are available in the Supplement material.^20^ Deduplication from downloaded references from databases was done using EndNote. As a complementary search: a search strategy was designed and used for Google Scholar, where the first 500 references were screened with no study selected for inclusion; citation searches on key studies were conducted.

Two independent reviewers (D.P., J.K.) screened the references in three phases (title/abstract, full-text, and detailed analysis during data extraction) according to the inclusion and exclusion criteria. Systematic process and blinded screening were performed using Rayyan^21^, and all disagreements were resolved by consensus.

### Assessment of risk of bias

For the assessment of study quality including the risk of bias and applicability concerns of included studies, the QUADAS-2 tool (Quality Assessment of Diagnostic Accuracy Studies)^22^ was used by two blinded investigators (D.P., G.S.). We excluded studies with a high risk of bias from our meta-analysis, whereas studies with an unclear risk of bias were marked and included in the analysis, along with studies labeled as low risk. The evaluation of noninvasive imaging (index test) of carotid plaques was blinded to histological evaluation (reference test) in studies.

### Data extraction

TP, TN, FP, and FN outcomes, and sensitivity and specificity were extracted from included studies by two independent readers (D.P., J.K.). The data extraction was performed according to the following criteria: when different techniques for plaque characteristic imaging were used in a study, we extracted data separately for each imaging modality; when the results of multiple reviewers were published in a study, we extracted one value as the sum of the results; when several different accuracy values were given based on cutoff points, the highest accuracy was extracted; when multiple plaque characteristics by imaging were compared to one plaque characteristic by pathology or one plaque characteristic by imaging compared to multiple plaque characteristics by histology, the finding with the highest accuracy was extracted to minimize the risk of bias in the calculation of the overall diagnostic accuracy of plaque characteristics; the study with the largest sample size was included to minimize duplicates or overlapping samples when authors published data from a single cohort or medical center more than once.

Disagreements between two readers were solved by consensus or by an independent decision of two senior team members (P.M., D.Š.) in all phases (reference screening, risk of bias, data extraction).

### Plaque instability and plaque characteristics

In *analysis 1*, we analyzed data of studies in which the investigators classified the plaques based on histology as vulnerable or stable according to their own definition, and evaluated the accuracy of MRI, CT, and/or US with regard to this histological classification.

In *analysis 2*, we first selected all studies in which specific histological plaque characteristics were assessed and studied the correlations of these plaque characteristics with MRI, CT, and/or US. Then we classified these studies into groups of vulnerable and stable plaque characteristics based on histology and evaluated the accuracy of imaging techniques. Vulnerable plaque characteristics were IPH (overall, acute/fresh, subacute/recent, old), LRNC, ruptured fibrous cap, ulceration, inflammation, and neovascularization, as in previous publications.^3–5,23^ In contrast, stable plaque characteristics were a predominant fibrous part, calcifications, intact fibrous cap, and loose matrix. If we found that some characteristic was alternatively considered to be vulnerable and stable in different published studies, we searched for more studies. For inclusion of the characteristic into one of the two groups, we had to find at least 80% studies indicating that the plaque characteristic was considered as stable, respective vulnerable. The assignment of a specific characteristics to one of the two groups also reflected the definitions used by investigators of the studies included in *analysis 1*.

### Statistics

Only studies with available data to calculate sensitivity and specificity and/or numbers of TP, TN, FP, and FN for any plaque characteristics were included in the meta-analysis. In case of incomplete data, corresponding or first authors were contacted to obtain the required information. In addition, we used formulas to compute TP, FP, FN, and TN values based on numbers of sensitivity, specificity, prevalence, positive predictive value, and negative predictive value.

First, we conducted separate univariate random-effects analyses of sensitivity and specificity to investigate the presence of heterogeneity in each index.^24^ Then, Rutter and Gatsonis’s hierarchical summary receiver operating characteristic (HSROC) curves were estimated for each imaging modality based on the bivariate random-effects model.^25–26^ The diagnostic performance (i.e. sensitivity and specificity) of the imaging modalities was assessed by comparing the HSROC curves. To facilitate the interpretation of results, the accuracy was also analyzed using univariate random-effects analyses based on the binomial distribution and logit transformation.

The pointwise confidence bands around the summary ROC curves were computed based on the multivariate delta method and tests of equality of the curves of Wald statistics. Studies reporting several pairs of sensitivity and specificity (because of the use of different imaging intensities) were considered independent given the sparsity of the data and convergence issues due to the complexity of the models. P-values < 0.05 were considered to indicate statistical significance.

The between-study variance τ^2^ was used to assess the heterogeneity. All the analyses were performed using Stata version 17 (Stata Corporation, College Station, TX, USA).

## Results

We identified 5960 studies by the literature search in four large databases. Supplementary searches were conducted in Google Scholar along with citation searching and five more studies were included. Finally, after the three-phase screening, 109 studies were found eligible. The details of the search strategy and study inclusion are presented in the PRISMA flow diagram in Figure 1 and in the Supplement.

**Figure 1.**
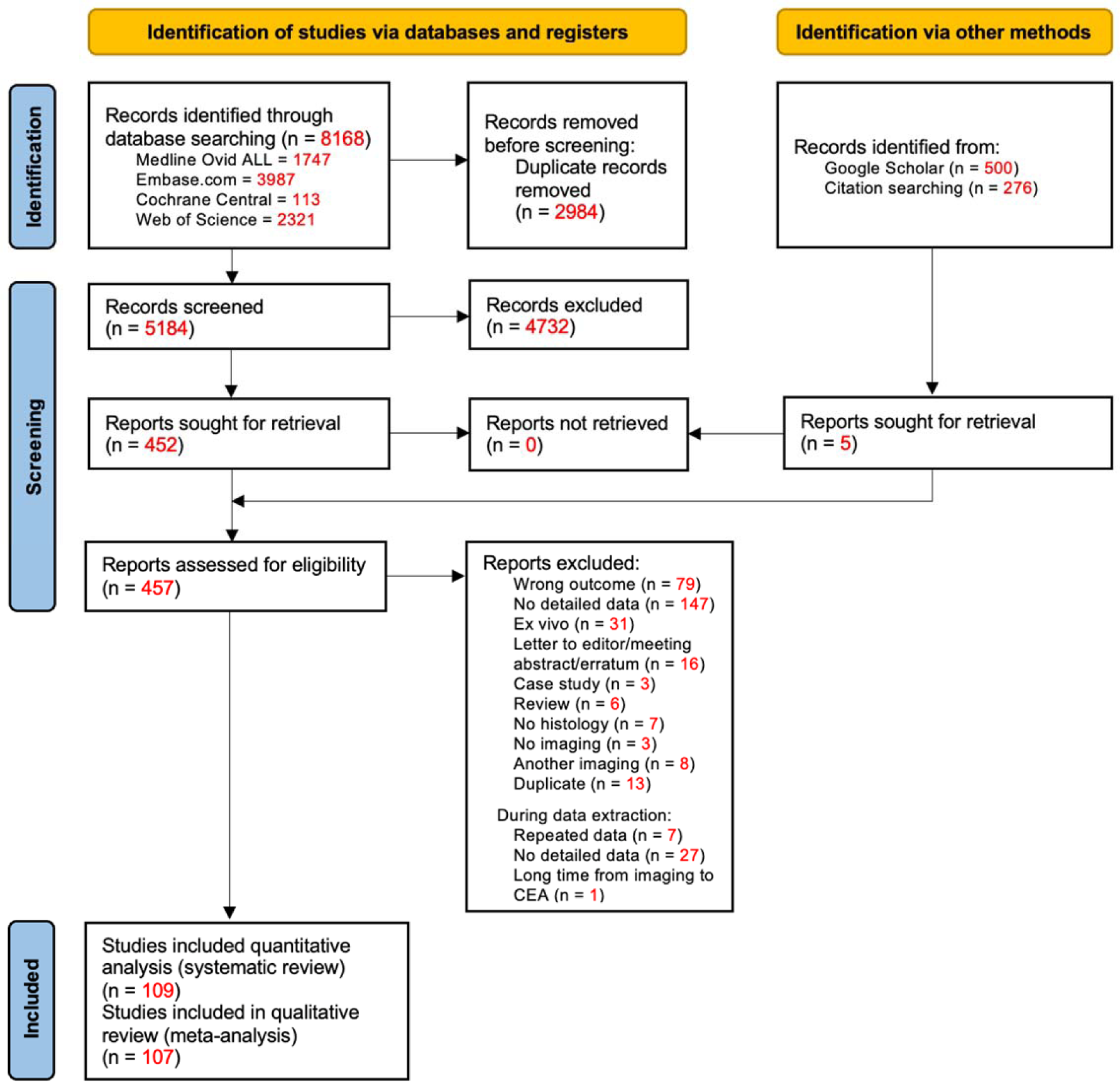
PRISMA flow chart diagram describing the publication search and selection algorithm.18

Before qualitative analysis, we evaluated the quality of the included studies and the possible risk of bias using QUADAS-2. Of the 109 included studies, we found two studies with a high risk of bias that we excluded. The overall results of the QUADAS-2 evaluation are presented in Figure 2 and detailed results are available in the Supplement (Data and Table S1).

**Figure 2.**
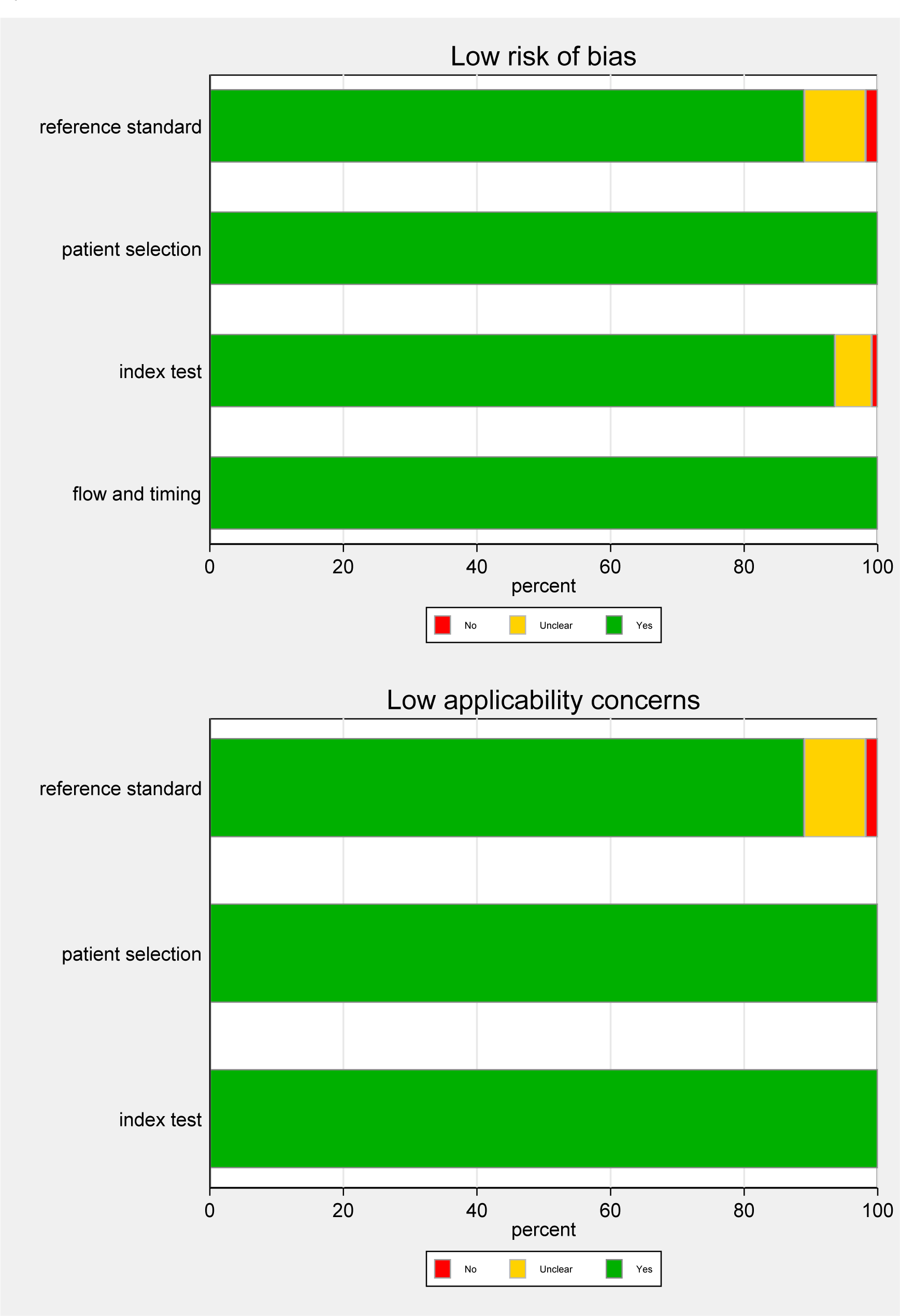
The risk of bias and the applicability concerns of included studies according to the QUADAS-2.22

For the qualitative analysis (meta-analysis) of the diagnostic accuracy in the included 107 studies, we found a total of 253 specific comparisons (28 CT, 120 MRI, and 105 US studies) of plaque instability and plaque characteristics from 6136 patients (Tables S2 and S3).

For *analysis 1* looking at classifying *plaques according to author-defined instability*, we identified 36 comparisons of vulnerable plaques and/or five comparisons of stable plaques in a total of 20 studies. The studies with the imaging technique taken into account, and definitions used for histological and imaging classification of plaques into vulnerable and stable are shown in Table 1, and the diagnostic accuracy of each comparison is shown in Figures 3, S3 and S4.

**Figure 3.**
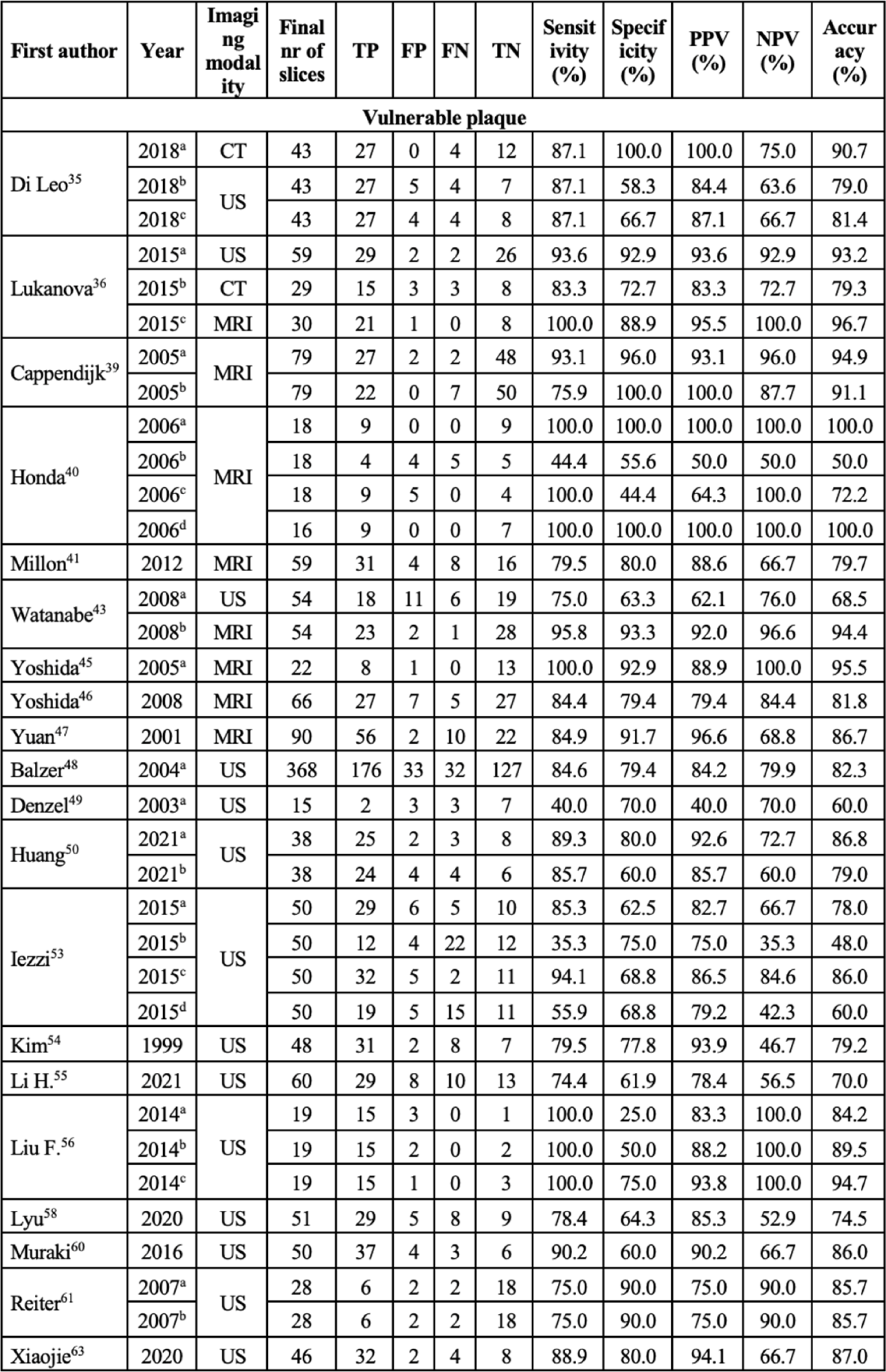
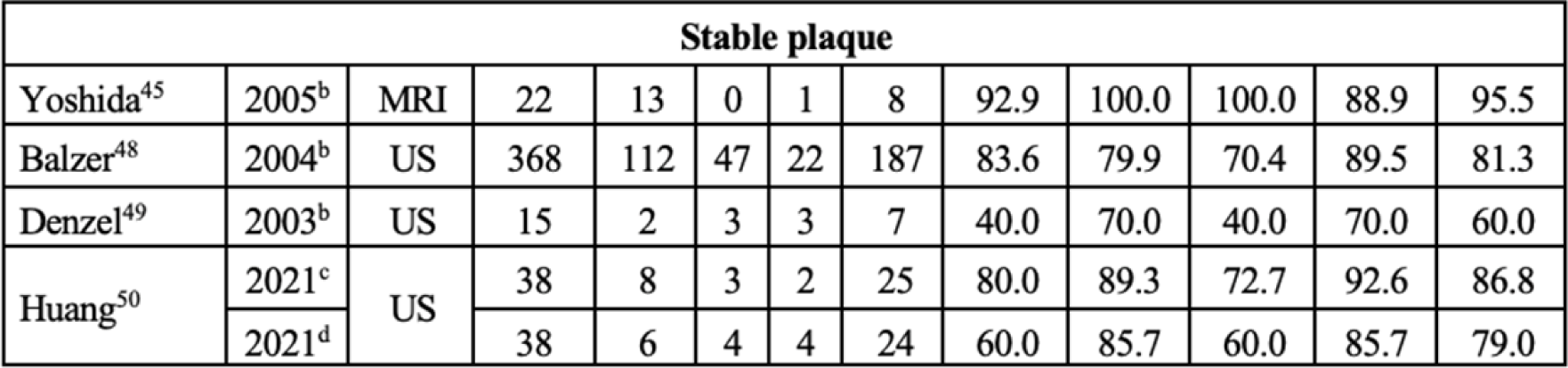
Diagnostic accuracy of 41 studies included in analysis 1 (A: vulnerable, B: stable).

**Table 1.** Studies (41 studies) identified for analysis 1 (“vulnerable” and “stable” plaques).

| First author | Year | Imaging | Details of imaging | Nr of patients | Plaque characteristic (imaging) | Plaque characteristic (histology) |
| --- | --- | --- | --- | --- | --- | --- |
| **Vulnerable plaque** |  |  |  |  |  |  |
| Di Leo<sup>35</sup> | 2018<sup>a</sup> | CT | native CT, CTA (130 ml) | 43 | negative HU (pre-contrast); >20 HU CE (post-contrast) | vulnerable plaque |
|  | 2018<sup>b</sup> | US | Doppler, CEUS |  | vulnerable plaque |  |
|  | 2018<sup>c</sup> |  | shear-wave elastography |  | vulnerable plaque (11-65 kPa) |  |
| Lukanova<sup>36</sup> | 2015<sup>a</sup> | US | duplex B-mode, color flow and power Doppler | 100 | unstable plaque: type I or II by Gray-Weale<sup>37</sup> | soft unstable plaque |
|  | 2015<sup>b</sup> | CT | native CT, CTA (80 ml) |  | unstable plaque (<149 HU) |  |
|  | 2015<sup>c</sup> | MRI | pre-/post-contrast T1w, ECG-triggered T2w + PDw |  | unstable plaque (AHA class IV-V and VI)<sup>38</sup> |  |
| Cappendijk<sup>39</sup> | 2005<sup>a</sup> | MRI | quantitative analysis | 11 | hemorrhage and lipid core/vulnerable plaque | vulnerable plaque |
|  | 2005<sup>b</sup> |  | semi-quantitative analysis |  |  |  |
| Honda<sup>40</sup> | 2006<sup>a</sup> | MRI | 2D TOF | 17 | high signal intensity | unstable plaque (AHA class ≥IV)<sup>38</sup> |
|  | 2006<sup>b</sup> |  | PDw |  |  |  |
|  | 2006<sup>c</sup> |  | T1w |  |  |  |
|  | 2006<sup>d</sup> |  | T2w |  |  |  |
| Millon<sup>41</sup> | 2012 | MRI | 3D TOF, T1w, PDw, CE-T1w | 69 | plaque CE | vulnerable plaque (AHA class VI or Lovett<sup>42</sup> grade 3-4) |
| Watanabe<sup>43</sup> | 2008<sup>a</sup> | US | GSM of B-mode, Doppler | 57 | at-risk soft plaque – type 1 or 2 by Geroulakos scale<sup>44</sup> | at-risk soft plaque |
|  | 2008<sup>b</sup> | MRI | 3D TOF, T1w, T2w |  | soft plaque |  |
| Yoshida<sup>45</sup> | 2005<sup>a</sup> | MRI | ECG-gated T1w + T2w | 26 | soft plaque | soft plaque |
| Yoshida<sup>46</sup> | 2008 | MRI | 3D GE, T1w, T2w | 70 | soft plaque (T1w cutoff: 1.25) | soft plaque |
| Yuan<sup>47</sup> | 2001 | MRI | 3D TOF, T1w, cardiac-gated PDw + T2w | 18 | soft plaque | soft plaque |
| Balzer<sup>48</sup> | 2004<sup>a</sup> | US | B-mode, | 368 | soft plaque | soft plaque |
|  |  |  | Doppler |  |  |  |
| Denzel<sup>49</sup> | 2003<sup>a</sup> | US | B-mode<br>GSM, duplex<br>US | 15 | GSM <35 | soft plaque |
| Huang<sup>50</sup> | 2021<sup>a</sup> | US | CEUS | 38 | vulnerable plaque – grade III–IV | vulnerable plaque<br>(presence of one<br>major or two minor<br>criteria of Naghavi<sup>52</sup>) |
|  | 2021<sup>b</sup> |  | 2D US |  | vulnerable plaque (score >4 by<br>Fabiani<sup>51</sup>) |  |
| Iezzi<sup>53</sup> | 2015<sup>a</sup> | US | low-dose<br>early-phase<br>(dynamic)<br>CEUS (2 ml) | 50 | enhancement flow | vulnerable (unstable)<br>plaque prone to<br>rupture |
|  | 2015<sup>b</sup> |  | low-dose<br>late-phase<br>(flash)<br>CEUS (2 ml) |  | enhancement |  |
|  | 2015<sup>c</sup> |  | high-dose<br>early-phase<br>(dynamic)<br>CEUS (4 ml) |  | enhancement flow |  |
|  | 2015<sup>d</sup> |  | high-dose<br>late-phase<br>(flash) CE<br>US (4 ml) |  | enhancement |  |
| Kim<sup>54</sup> | 1999 | US | duplex,<br>Doppler | 65 | echolucent plaque | soft plaque |
| Li H.<sup>55</sup> | 2021 | US | GSM of US,<br>color/pulsed<br>Doppler, CE<br>US | 124 | enhancement (grade II) | vulnerable plaque |
| Liu F.<sup>56</sup> | 2014<sup>a</sup> | US | B-mode | 19 | vulnerable plaque | vulnerable plaque |
|  | 2014<sup>b</sup> |  | real-time<br>elastography |  | vulnerable plaque (score of 1-3<br>on Itoh scale)<sup>57</sup> |  |
|  | 2014<sup>c</sup> |  | B-mode +<br>real-time<br>elastography |  | vulnerable plaque |  |
| Lyu<sup>58</sup> | 2020 | US | standard US,<br>CEUS | 70 | enhancement (grade 2–3) | vulnerable plaque<br>(Oxford plaque<br>study<sup>59</sup> grade 3–4) |
| Muraki<sup>60</sup> | 2016 | US | standard US | 50 | fine trembling motion of<br>echogenic structures inside the<br>plaque | soft content within<br>plaque |
| Reiter<sup>61</sup> | 2007<sup>a</sup> | US | B-mode | 28 | soft plaque by Beletsky<sup>62</sup>: group<br>1 | soft plaque/organized<br>thrombus by<br>Beletsky<sup>62</sup> |
|  | 2007<sup>b</sup> |  | B-flow<br>imaging |  |  |  |
| Xiaojie<sup>63</sup> | 2020 | US | Doppler | 44 | vulnerable plaque (score >4 by<br>Fabiani<sup>51</sup>) | vulnerable plaque<br>(presence of one<br>major or two minor<br>criteria of Naghavi<sup>52</sup>) |
| **Stable plaque** |  |  |  |  |  |  |
| Yoshida<sup>45</sup> | 2005<sup>b</sup> | MRI | ECG-gated<br>T1w + T2w | 26 | hard plaque | hard plaque |
| Balzer<sup>48</sup> | 2004<sup>b</sup> | US | B-mode,<br>Doppler | 368 | hard plaque | hard plaque |
| Denzel<sup>49</sup> | 2003<sup>b</sup> | US | B-mode, | 15 | GSM >65 | hard plaque (calcium- |
|  |  |  | duplex US,<br>GSM |  |  | rich) |
| Huang<sup>50</sup> | 2021<sup>c</sup> | US | CE US | 38 | stable plaque – grade I–II | stable plaque<br>(absence of one<br>major or two minor<br>criteria of Naghavi<sup>52</sup>) |
| | 2021<sup>d</sup> | | 2D US | | stable plaque (score $\leq 4$ by<br>Fabiani<sup>51</sup>) | |

CE – contrast enhanced/enhancement; ECG – echocardiography; GE – gradient echo; GSM – Gray Scale Median; HU – Hounsfield unit; IPH – intraplaque hemorrhage; TOF – time of flight, PD – proton dense

For *analysis 2* assessing *several plaque characteristics*, the 212 remaining comparisons were divided into vulnerable (169) and stable (42) plaques as depicted in Table S3. One characteristic with insufficient details (mixed plaque) was excluded. Details of all evaluated characteristics by studies are available in Table S2. Most of the studies evaluated multiple characteristics.

### Univariate analyses

In *analysis 1 (plaque instability according to the author’s definition)*, we found that MRI had a very high diagnostic accuracy in visualizing vulnerable plaque, with a pooled accuracy of 90%, followed by CT (86%) and US (80%) as presented in Figure 4 and Table 2. Regarding the imaging of stable plaques, a very high accuracy was noted in one identified MRI study (95%). No CT-based study was found, and four US-based studies yielded an accuracy of 81% (Figure S5, Table 2).

**Figure 4.**
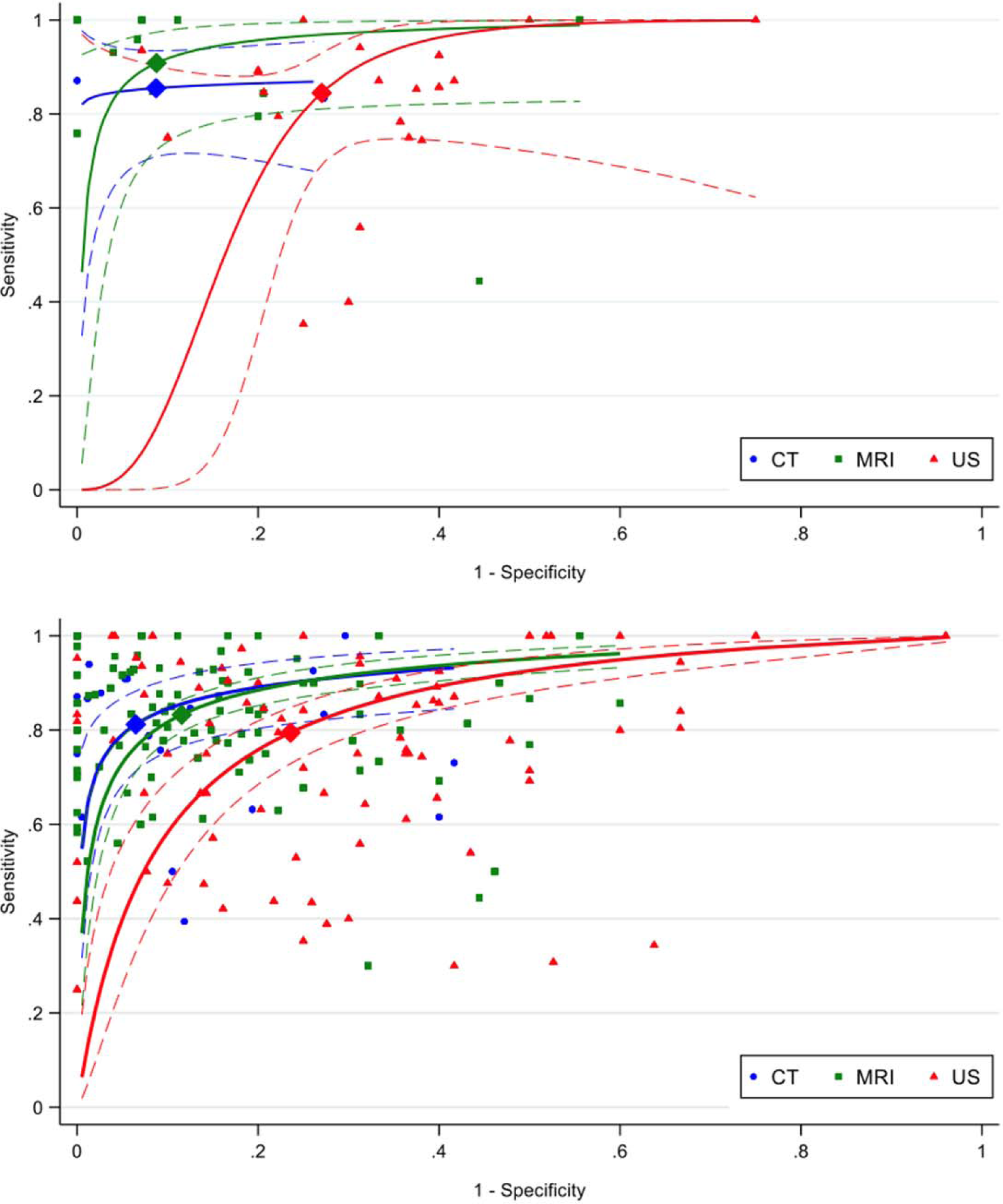
HSROC curves of analysis 1 (top) and analysis 2 (bottom) for visualization of vulnerable plaque and vulnerable plaque characteristics, respectively, based on HSROC curves.

**Table 2.** Meta-analysis of diagnostic accuracy of vulnerable and stable plaque (analysis 1), and of vulnerable and stable plaque characteristics (analysis 2).

| Plaque features | CT |  |  | MRI |  |  | US |  |  |
| --- | --- | --- | --- | --- | --- | --- | --- | --- | --- |
| | Sensitivity %<br>(95% CI)<br>$\tau^2$ | Specificity %<br>(95% CI)<br>$\tau^2$ | Accuracy %<br>(95% CI)<br>$\tau^2$ | Sensitivity %<br>(95% CI)<br>$\tau^2$ | Specificity %<br>(95% CI)<br>$\tau^2$ | Accuracy %<br>(95% CI)<br>$\tau^2$ | Sensitivity %<br>(95% CI)<br>$\tau^2$ | Specificity %<br>(95% CI)<br>$\tau^2$ | Accuracy %<br>(95% CI)<br>$\tau^2$ |
|  | Author-defined plaque instability (Analysis 1) |  |  |  |  |  |  |  |  |
| Vulnerable plaque | 86<br>(73-93)<br>0.00 | 87<br>(66-96)<br>0.00 | 86<br>(76-92)<br>0.00 | 91<br>(80-96)<br>1.21 | 91<br>(80-96)<br>1.58 | 90<br>(82-95)<br>0.91 | 84<br>(78-90)<br>0.77 | 73<br>(67-78)<br>0.11 | 80<br>(75-84)<br>0.28 |
| Stable plaque | NA |  |  | 93*<br>(69-99)<br>NA | 100*<br>(68-100)<br>NA | 95*<br>(78-99)<br>NA | 75<br>(55-88)<br>0.21 | 81<br>(76-85)<br>0.00 | 81<br>(77-84)<br>0.00 |
| Several plaque characteristics (Analysis 2) |  |  |  |  |  |  |  |  |  |
| Vulnerable plaque characteristics | 81<br>(74-87)<br>0.58 | 94<br>(87-97)<br>2.37 | 89<br>(84-92)<br>0.72 | 83<br>(81-86)<br>0.41 | 88<br>(86-91)<br>0.90 | 86<br>(84-88)<br>0.39 | 80<br>(75-84)<br>1.14 | 76<br>(71-81)<br>1.15 | 77<br>(74-81)<br>0.66 |
| Stable plaque characteristics | 87<br>(74-97)<br>0.37 | 93<br>(63-99)<br>5.32 | 90<br>(66-98)<br>3.30 | 83<br>(79-87)<br>0.05 | 92<br>(86-96)<br>1.25 | 89<br>(85-92)<br>0.46 | 75<br>(67-82)<br>0.38 | 86<br>(78-91)<br>0.92 | 82<br>(75-88)<br>0.71 |

\* = Wilson 95% CI based on results from only one study; NA = not applicable; CI = confidence interval;
$\tau^2$ = between-study variance

In *analysis 2 (several plaque characteristics)*, CT had the highest accuracy in the detection of vulnerable and stable carotid plaque characteristics (89% and 90%, respectively). However, the results with CT showed a wider range of the 95% confidence interval (CI) (84– 92%) for vulnerable plaque characteristics compared to MRI (84–88%) and even for stable plaque characteristics (66–98% with CT vs 85–92% with MRI). CT and MRI had similar accuracy for vulnerable (89% and 86%, respectively) and stable (90% and 89%, respectively) plaque characteristics. Findings with the US modality showed the lowest sensitivity and specificity of all studied modalities in the diagnosis of vulnerable (specificity 79% and specificity 76%) and stable plaque characteristics (specificity 77% and specificity 86%). Detailed data are available in Table 2.

### Bivariate analyses

For *analysis 1* regarding vulnerable plaque, Figure 4 (top) shows that CT and MRI always outperform US by a higher specificity for a given sensitivity: the overall comparison of the three curves resulted in p=0.013, between CT and MRI in p=0.907, between CT and US in p=0.432, and between MRI and US in p=0.003. For stable plaque (Figure S5, top), it was only possible to estimate an HSROC curve for the US modality.

For *analysis 2*, Figure 4 (bottom) shows that CT and MRI again were the most accurate imaging techniques for detecting vulnerable plaque characteristics, with the US modality always showing the lowest specificity. The comparison between the three curves resulted in p<0.001, between CT and MRI in p=0.201, between CT and US in p<0.001, and between MRI and US in p<0.001. For stable plaque characteristics (Figure S5, bottom), the comparison between the three curves showed p=0.146, between CT and MRI p=0.812, between CT and US p=0.287, and between MRI and US p=0.054. This is in line with the results regarding the accuracy parameters provided in Table 2.

Finally, Figures S6–S8 show that there was large heterogeneity among the studies for all three imaging techniques.

## Discussion

Our study showed that the CT imaging technique had the highest accuracy for detecting vulnerable plaque and vulnerable plaque characteristics, not statistically significant, and the MRI was able to identify all vulnerable plaque characteristics with similar accuracy as CT. Regarding the stable plaque characteristics, CT and MRI showed almost the same accuracy (90% vs 89%, respectively). US proved to have significantly lower performance for detection of vulnerable plaque/plaque characteristics and nonsignificantly lower diagnostic accuracy for detection of stable plaque characteristics.

Although some studies have suggested that CT performs less well in identifying high-risk plaque characteristics,^27^ in our study CT showed good diagnostic accuracy in visualizing both vulnerable and stable plaque. Previous systematic reviews confirmed that CT is an accurate noninvasive modality in atherosclerotic lesions evaluation.^28^ However, compared to MRI, CT failed to detect all vulnerable components, but the most common at-risk plaque characteristics (IPH, LRNC, or ulceration) could be identified. CT may represent a good choice in carotid plaque diagnostics due to its excellent spatial resolution, precision, and widespread availability and short examination time.^29^ It also has been demonstrated that the identification of vulnerable characteristics using CT can predict 10-year atherosclerotic cardiovascular disease risk.^30–31^ According to our results, CT is an accurate diagnostic modality to evaluate vulnerable or stable plaque with an accuracy of 89% and 90%, respectively.

MRI demonstrated the highest performance with good diagnostic accuracy for most plaque characteristics in our study and a previous one.^16^ Imaging of carotid arteries by MRI is an established, fast, reliable, and accurate diagnostic modality, which has many advantages over other imaging techniques. Most of the known vulnerable and stable plaque characteristics can be visualized using MRI.^32^ The results of our meta-analysis confirmed these previously reported results because MRI visualized all 13 vulnerable and stable plaque characteristics evaluated in the included studies (compared to 5 using CT and 9 using US). It was proven that MRI is a good accurate noninvasive imaging modality for vulnerable plaque diagnosis (accuracy of 90% compared to 86% of CT and 80% of US). We could not assess the performance of MRI for the detection of stable plaques as there was only one MRI study that evaluated stable plaque (sensitivity 93%, specificity 100%). It was demonstrated that CT was a useful modality to visualize both vulnerable and stable plaque characteristics.

Imaging by the US technique is widely employed in clinical practice and often represents the first choice of carotid artery stenosis examination, because of low cost, its repeatability and there are no contraindications.^33^ Therefore, US is recommended as a screening modality for carotid atherosclerosis.^34^ In our study, US was the noninvasive imaging modality with less diagnostic accuracy for stable or vulnerable carotid plaque features. Nevertheless, the accuracy compared to histology as the reference standard was still high (over 75%). US could be used as a screening and first-line modality to assess carotid plaque instability or plaque progression.

There were several limitations that must be taken into account. First, we did not analyze different imaging techniques separately (e.g. non non-contrast US and contrast-enhanced US; MRI sequences) or different used thresholds separately which may represent a potential bias. However, the objective of the meta-analysis was to provide the overall diagnostic accuracy of noninvasive imaging modalities, regardless of differences between techniques or cutoff points. Second, the accuracy of imaging modalities with better resolution increased over time due to technical progress. Therefore, older included studies may have influenced the results of the overall accuracy of imaging modalities. Third, the publication date of the included studies in the analysis was not restricted. Although this point could have led to a potential bias due to advances in imaging technology, Figures S1 and S2 showed that the diagnostic accuracy of the studies included in the stable or vulnerable plaque characteristics group not changed clearly over time. Finally, a few studies were available for certain characteristics, therefore the diagnostic accuracy may be less reliable.

## Conclusion

In our study, it was proven that CT and MRI were good noninvasive modalities for detection of vulnerable plaques with similar, high diagnostic accuracy. However, all vulnerable plaque characteristics were visualized with MRI leading to a comprehensive assessment of plaque instability. The US technique showed significantly less accuracy and could be considered for baseline screening and follow-up. CT and MRI should be used to better identify plaque instability and guide management to reduce the stroke risk.

## Nonstandard Abbreviations

IPH: intraplaque hemorrhage
LRNC: lipid-rich necrotic core
US: ultrasound
PRISMA: Preferred Reporting Items for Systematic Reviews and Meta-analyses
STARD: Standards for Reporting of Diagnostic Accuracy Studies
QUADAS: Quality Assessment of Diagnostic Accuracy Studies
HSROC: hierarchical summary receiver operating characteristic

## Data Availability

Detailed statistical analyses, major resources tables and figures can be found in the Supplemental Material. All material, data, and detailed protocols are available upon request.

## Acknowledgment

We are very thankful to Aristeidis H. Katsanos, Karen Von Deneen, Di Dong, Yukari Yamada, Šárka Kostecká and Cecile Jaques for their help with our study.

## Sources of funding

None related to the research, authorship, and/or publication of this article. David Pakizer was supported by VIA PhD grant University of Ostrava for travel related to this analysis. Statistical analysis was supported by grants NV19-04-00270, NU22-04-00389, SGS11/LF/2022; and by University of Lausanne, Faculty of Biology and Medicine.

## Disclosures

None.

**Supplemental material**

Supplemental Statistics Supplemental Results Tables S1–S3

## Notes

### Competing Interest Statement

The authors have declared no competing interest.

### Clinical Trial

PROSPERO ID CRD42022329690 (https://www.crd.york.ac.uk/prospero/display_record.php?RecordID=329690)

